# mRNA vaccination compared to infection elicits an IgG-predominant response with greater SARS-CoV-2 specificity and similar decrease in variant spike recognition

**DOI:** 10.1101/2021.04.05.21254952

**Authors:** Katharina Röltgen, Sandra C. A. Nielsen, Prabhu S. Arunachalam, Fan Yang, Ramona A. Hoh, Oliver F. Wirz, Alexandra S. Lee, Fei Gao, Vamsee Mallajosyula, Chunfeng Li, Emily Haraguchi, Massa J. Shoura, James L. Wilbur, Jacob N. Wohlstadter, Mark M. Davis, Benjamin A. Pinsky, George B. Sigal, Bali Pulendran, Kari C. Nadeau, Scott D. Boyd

**Affiliations:** Department of Pathology, Stanford University School of Medicine, Stanford, CA, USA; Institute for Immunity, Transplantation and Infection, Stanford University, Stanford, CA, USA; Sean N. Parker Center for Allergy & Asthma Research, Stanford, CA, USA; Meso Scale Diagnostics LLC, Rockville, Maryland, USA; Department of Microbiology and Immunology, Stanford University, Stanford, CA, USA; Howard Hughes Medical Institute, Stanford University, Stanford, CA, USA; Department of Medicine, Division of Infectious Diseases and Geographic Medicine, Stanford University, Stanford, CA USA; Department of Medicine, Division of Pulmonary, Allergy & Critical Care Medicine, Stanford University, Stanford, CA, USA

**Keywords:** COVID-19, BioNTech/Pfizer BNT162b2, mRNA vaccine, serology, electrochemiluminescence, SARS-CoV-2, variants of concern, endemic coronaviruses, antibodies

## Abstract

During the severe acute respiratory syndrome coronavirus 2 (SARS-CoV-2) pandemic, new vaccine strategies including lipid nanoparticle delivery of antigen encoding RNA have been deployed globally. The BioNTech/Pfizer mRNA vaccine BNT162b2 encoding SARS-CoV-2 spike protein shows 95% efficacy in preventing disease, but it is unclear how the antibody responses to vaccination differ from those generated by infection. Here we compare the magnitude and breadth of antibodies targeting SARS-CoV-2, SARS-CoV-2 variants of concern, and endemic coronaviruses, in vaccinees and infected patients. We find that vaccination differs from infection in the dominance of IgG over IgM and IgA responses, with IgG reaching levels similar to those of severely ill COVID-19 patients and shows decreased breadth of the antibody response targeting endemic coronaviruses. Viral variants of concern from B.1.1.7 to P.1 to B.1.351 form a remarkably consistent hierarchy of progressively decreasing antibody recognition by both vaccinees and infected patients exposed to Wuhan-Hu-1 antigens.

## Introduction

In 2020, following decades of research to develop messenger RNA (mRNA) vaccines, and accelerated by the urgent need for countermeasures against the coronavirus disease 2019 (COVID-19) pandemic, the U.S. FDA authorized two mRNA vaccines, BNT162b2 (BioNTech/Pfizer) and mRNA-1273 (Moderna/NIAID). mRNA vaccines are a promising alternative to conventional vaccine approaches in part because a relatively consistent biomolecule can be used to generate a variety of antigens in the vaccine recipient (Pardi et al., 2018a). They have been shown to stimulate protective immune responses to viral infections in pre-clinical models (Pardi et al., 2017, 2019; Richner et al., 2017; Vogel et al., 2021), and recently have demonstrated high efficacy and safety in clinical trials for COVID-19 prevention (Baden et al., 2021; Polack et al., 2020; Walsh et al., 2020). mRNA vaccines mimic some aspects of viral infection by using the host cell’s translational machinery to transiently express properly folded vaccine antigens *in situ*, driving strong humoral and T cell responses (Sahin et al., 2014; Zhang et al., 2019). It remains to be determined precisely how the immune system responds to RNA vaccines and their other components such as lipid nanoparticles, compared to other vaccine platforms or infection.

Immune correlates of protection from COVID-19 have not been fully elucidated, but both humoral and cellular responses may contribute to preventing and containing infection (McMahan et al., 2021; Ni et al., 2020; Rydyznski Moderbacher et al., 2020). Most neutralizing antibodies target the receptor-binding domain (RBD) of SARS-CoV-2 spike (S) and prevent binding to the host angiotensin-converting enzyme 2 (ACE2) receptor (Yuan et al., 2021). The BNT162b2 mRNA vaccine (as well as mRNA-1273) encodes full-length prefusion stabilized S glycoprotein. Results from Phase III clinical trials and mass vaccination studies show promising results with high efficacy against severe COVID-19 and similar data across subgroups defined by age, sex, race, and the presence of coexisting conditions (Dagan et al., 2021; Polack et al., 2020). BNT162b2 elicited robust anti-S IgG responses and SARS-CoV-2 neutralizing titers in the trials (Walsh et al., 2020). The emergence and global spread of SARS-CoV-2 variants of concern with mutations in the S gene first detected in the United Kingdom (B.1.1.7 lineage), South Africa (B.1.351 lineage), and Brazil (P.1 lineage), threaten to decrease the efficacy of vaccines based on the original Wuhan-Hu-1 SARS-CoV-2 S antigen. All three variants have a N501Y amino acid change in RBD, while the B.1.351 and P.1 variants both have two additional RBD changes, K417N/T and E484K, increasing the binding affinity of RBD to ACE2 (Ramanathan et al., 2021). These amino acid changes, particularly E484K, alter important epitopes targeted by many antibodies that neutralize SARS-CoV-2 by preventing RBD binding to host ACE2 (Greaney et al., 2021).

Here, we compare the longitudinal antibody responses in 55 BNT162b2 vaccine recipients and 100 COVID-19 patients, and identify key differences in the magnitude, isotype profiles, SARS-CoV-2 S domain specificity and breadth of responses targeting other human coronaviruses (HCoVs). In contrast, evaluating IgG and RBD-ACE2 blocking antibody responses to the early Wuhan-Hu-1 S protein and the three most concerning novel viral variants B.1.1.7, P.1 and B.1.351, we find remarkably consistent vulnerabilities among different individuals regardless of whether their antibody responses were stimulated by infection or vaccination.

## Results

### BNT162b2 vaccination induces high anti-SARS-CoV-2 IgG concentrations

We measured anti-SARS-CoV-2 antibody isotype concentrations for nucleocapsid (N), full S and S domains S1 N-terminal domain (NTD) and RBD in BNT162b2 study participant plasma samples collected before or immediately after their first dose (day 0), and on days 7, 21 (prior to the 2^nd^ dose), 28, and 42 after the prime using multiplexed electrochemiluminescence (ECL) assays (Meso Scale Discovery, MSD). Four of the vaccine recipients had a history of positive SARS-CoV-2 RT-qPCR tests (CoV-2+ vaccinees), while the remaining 51 participants were naïve to SARS-CoV-2 (CoV-2– vaccinees). IgG titers for S protein and its domains in CoV-2– vaccinees were negative at baseline and day 7 after their first vaccination, increased by day 21, and reached their highest levels at days 28 and 42 (Figures 1A and 1B). IgG to N protein in CoV-2– participants remained negative throughout the vaccine course as expected. The kinetics of the S protein IgG response tended to be faster for patients previously infected with SARS-CoV-2. Three of the four CoV-2+ vaccinees showed elevations in IgG antibody responses to NTD, RBD, and S by day 7 and reached higher levels than observed for the CoV-2– vaccinees at day 21. One CoV-2+ vaccinee showed slower kinetics of IgG increase, more similar to CoV-2– vaccinees (Figures 1A and 1B). CoV-2+ participant median anti-N protein IgG concentrations were increased at baseline and did not increase with vaccination.

**Figure 1.**
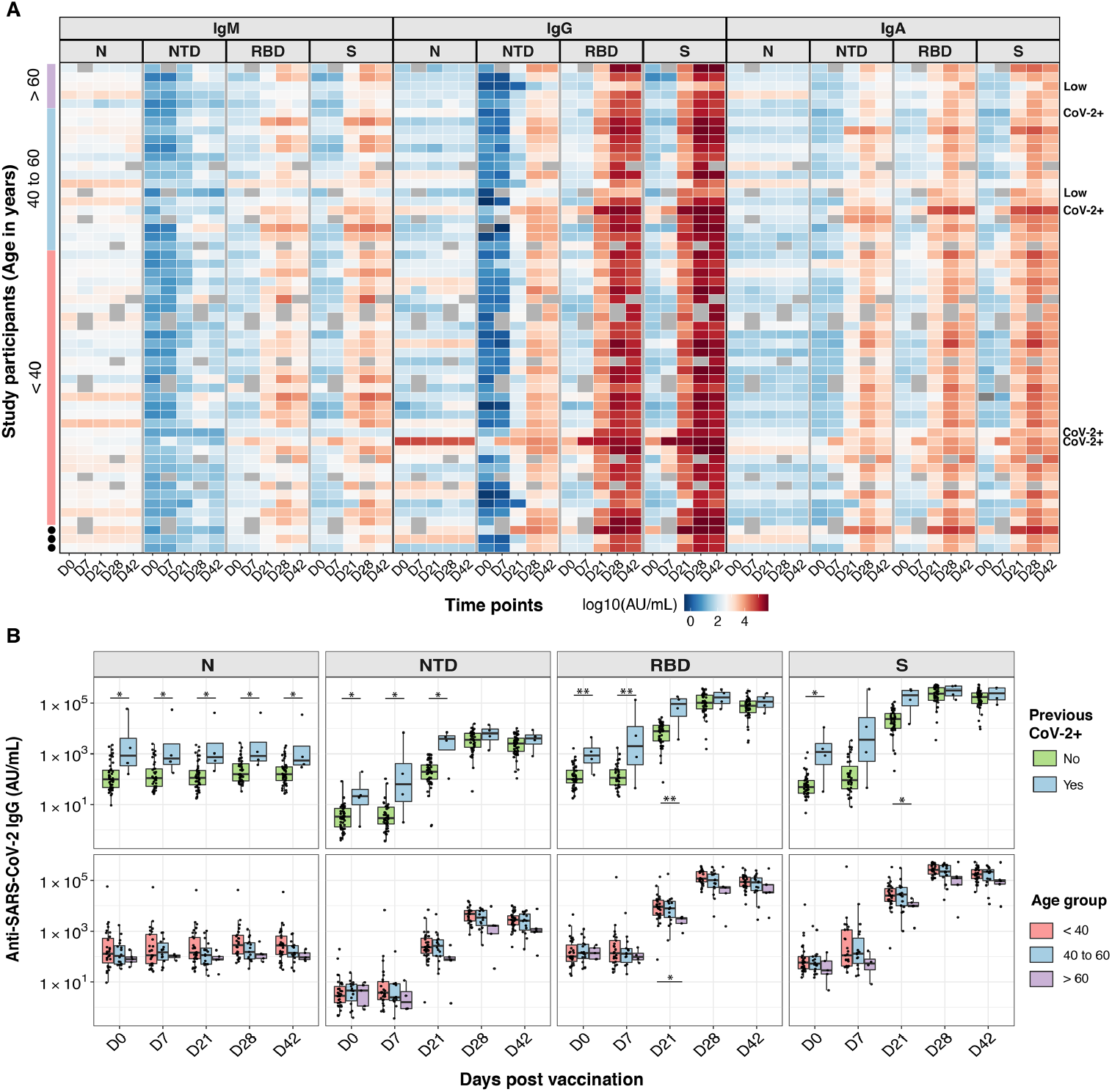
BNT162b2 vaccination induces high anti-SARS-CoV-2 IgG concentrations. Anti-SARS-CoV-2 N, NTD, RBD, and S IgM, IgG, and IgA antibody responses are shown for 257 plasma samples from 55 individuals who received BNT162b2 prime (day 0) and boost (day 21) vaccination doses. (A) Heatmap showing the development of antibody responses in longitudinal samples collected at day 0, 7, 21, 28, and 42 post-prime vaccination (x-axis). Log10 MSD arbitrary unit (AU)/mL concentrations are displayed for study participants sorted by age (y-axis, color-coded). Rows are labeled on the right with “CoV-2+” for participants with a previous SARS-CoV-2 RT-qPCR positive test result and with a “Low” for participants with low antibody concentrations at day 28 and day 42 post-prime. (B) Box-whisker plots of the MSD AU/mL anti-SARS-CoV-2 IgG concentrations show the interquartile range as the box and the minimum and maximum values as the ends of the whiskers. Comparisons between two groups (CoV-2+ and CoV-2) were by the two-sided Wilcoxon rank sum test; comparison between age groups (< 40; 40 to 60; > 60 years) was tested using pairwise Wilcoxon rank sum test with Bonferroni correction. * = P < 0.05, ** = P < 0.01, *** = P < 0.001.

All but two study participants had developed robust IgG responses to SARS-CoV-2 RBD and S by day 28 (Figure 1A), with CoV-2+ and CoV-2– vaccinees reaching comparable IgG concentrations (Figures 1A and 1B). One individual with low anti-RBD and anti-S responses did not receive the second vaccine dose; the other was a 70-year-old individual with a prior history of oral cancer. Two vaccinees taking methotrexate immunosuppressive medication showed no decrease in antibody responses to vaccination. All BNT162b2 recipients had weaker IgM and IgA antibody responses to S domains and full S, in comparison to their IgG responses (Figure 1A; Figures S1A and S1B).

Study participants in all age groups (< 40 years, 40 to 60 years, > 60 years) developed robust IgG antibody responses, although younger individuals reached higher Ig antibody concentrations compared to the individuals over 60 years of age (Figures 1A and 1B; Figures S1A and S1B). We compared IgG antibody responses in vaccinees who did or did not report side effects from their prime and boost vaccination. The most common side effects reported were site tenderness, muscle aches, headaches, and fatigue (Figure S2A). None of the side effects experienced after the prime or boost vaccination were associated with an increase or decrease in IgG antibody responses (Figure S2B).

### BNT162b2 vaccination and SARS-CoV-2 infection elicit distinct Ig isotype profiles

COVID-19 patients with severe disease develop higher SARS-CoV-2-specific antibody titers than asymptomatic or mildly ill individuals (Long et al., 2020; Röltgen et al., 2020). We measured the magnitude and Ig isotype profiles in moderately and severely ill COVID-19 patients sampled in the initial months of the pandemic before viral variants of concern had been reported, and compared these to the responses of BNT162b2 vaccinees, using the MSD ECL platform (Figure 2; Figure S3).

**Figure 2.**
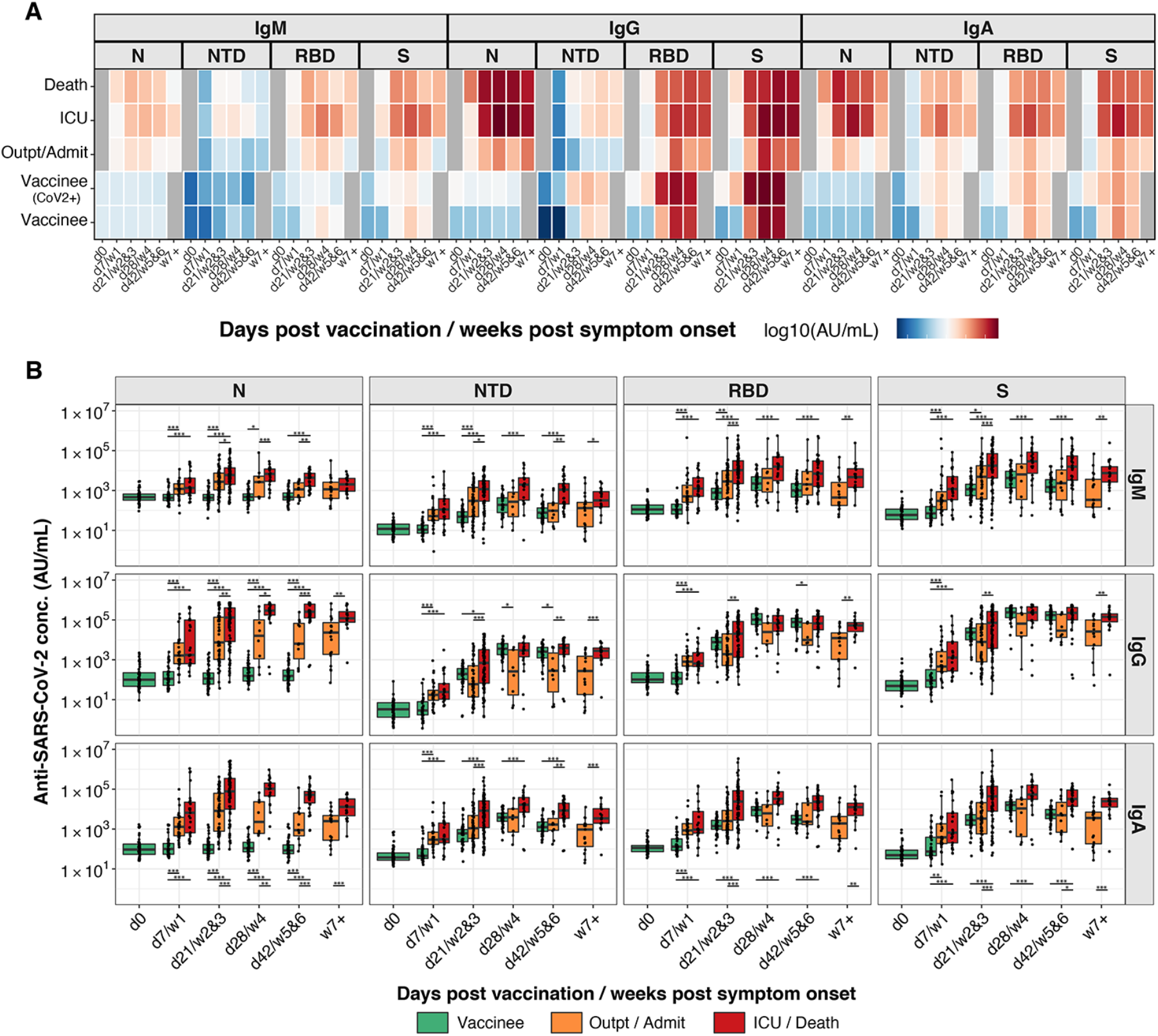
BNT162b2 vaccination and SARS-CoV-2 infection elicit distinct Ig isotype profiles. Anti-SARS-CoV-2 N, NTD, RBD, and S IgM, IgG, and IgA antibody responses are shown for individuals who received BNT162b2 prime (day 0) and boost (day 21) vaccination doses and for COVID-19 patients. (A) Heatmap showing the development of antibody responses in longitudinal samples from vaccinees/patients collected at / during day 0, day 7 / week 1, day 21 / weeks 2&3, day 28 / week 4, day 42 / weeks 5&6, and week 7 and later after vaccination / COVID-19 symptom onset (x-axis). Individuals were classified as outpatients (Outpt) and hospital admitted patients (Admit); intensive care unit (ICU) patients, those who died from their illness (Death) and vaccinees who did (CoV-2+) or did not have a positive SARS-CoV-2 test in the past. The color scale encodes the median values of log10 MSD arbitrary unit (AU)/mL concentrations. (B) Box-whisker plots of the MSD AU/mL anti-SARS-CoV-2 Ig concentrations show the interquartile range as the box and the minimum and maximum values as the ends of the whiskers. Statistical test for significance between groups (Outpatient/Admit; ICU/Death; Vaccinees) was performed using pairwise Wilcoxon rank sum test with Bonferroni correction. * = P < 0.05, ** = P < 0.01, *** = P < 0.001.

COVID-19 plasma samples were from a previously described cohort of patients who presented to Stanford Healthcare clinical sites for care (Röltgen et al., 2020). Patients were classified as outpatients; admitted patients not requiring care in the intensive care unit (ICU); ICU patients; and those who died from their illness. Serological responses measured by ECL in 530 longitudinal plasma samples from these 100 COVID-19 patients were highly correlated with results from laboratory-developed anti-RBD, -S1, and -N ELISAs (Figure S4) (Röltgen et al., 2020).

Vaccinees developed IgG antibody concentrations to the SARS-CoV-2 NTD, RBD, and S proteins that were comparable to the responses in severely ill patients, and higher than those of mildly or moderately ill patients; this reached statistical significance for anti-NTD antibodies at days 28 and 42 and for anti-RBD antibodies at day 42. In comparison to infection, the BNT162b2 vaccine induced a highly IgG-polarized serological response, with minimal IgM and IgA responses to S and S domains RBD and NTD (Figure 2; Figure S3).

### BNT162b2 vaccination produces less broad serological responses to endemic HCoVs than SARS-CoV-2 infection

While SARS-CoV-2 proteins show sequence divergence from those of other HCoVs, regions of high conservation exist at the epitope level (Ladner et al., 2021) that can lead to plasma antibody cross-reactivity. SARS-CoV-2 and SARS-CoV S proteins share 76% amino acid similarity. Serological analysis cannot readily distinguish between antibodies produced after reactivation of pre-existing HCoV antigen-specific memory B cells by SARS-CoV-2 antigens, or stimulation of novel cross-reactive antibody species by SARS-CoV-2 vaccination or infection.

In SARS-CoV-2 vaccinees and COVID-19 patients within the first weeks after vaccination or infection, respectively, antibody responses to SARS-CoV S increased. Vaccinees and severely ill patients developed similar concentrations of anti-SARS-CoV S IgG, whereas those of moderately ill patients were significantly lower. Severely ill patients had higher stimulation of SARS-CoV anti-S IgA concentrations than vaccinees and other patients (Figures 3A and 3B; Figure S5). Infection stimulated notably higher IgG responses to betacoronaviruses HCoV-OC43 and HCoV-HKU1 compared to vaccination, despite the similar magnitude of total anti-SARS-CoV-2-specific antibody responses in vaccinees and severely ill patients (Figure 2B). IgA and IgM showed similar patterns of higher responses to endemic HCoVs in severely ill patients compared to vaccinees. HCoV responses stimulated by infection varied by isotype; notably, an IgA response to alphacoronavirus HCoV-NL63 S was seen consistently in the patient groups but was not present in vaccinated individuals.

**Figure 3.**
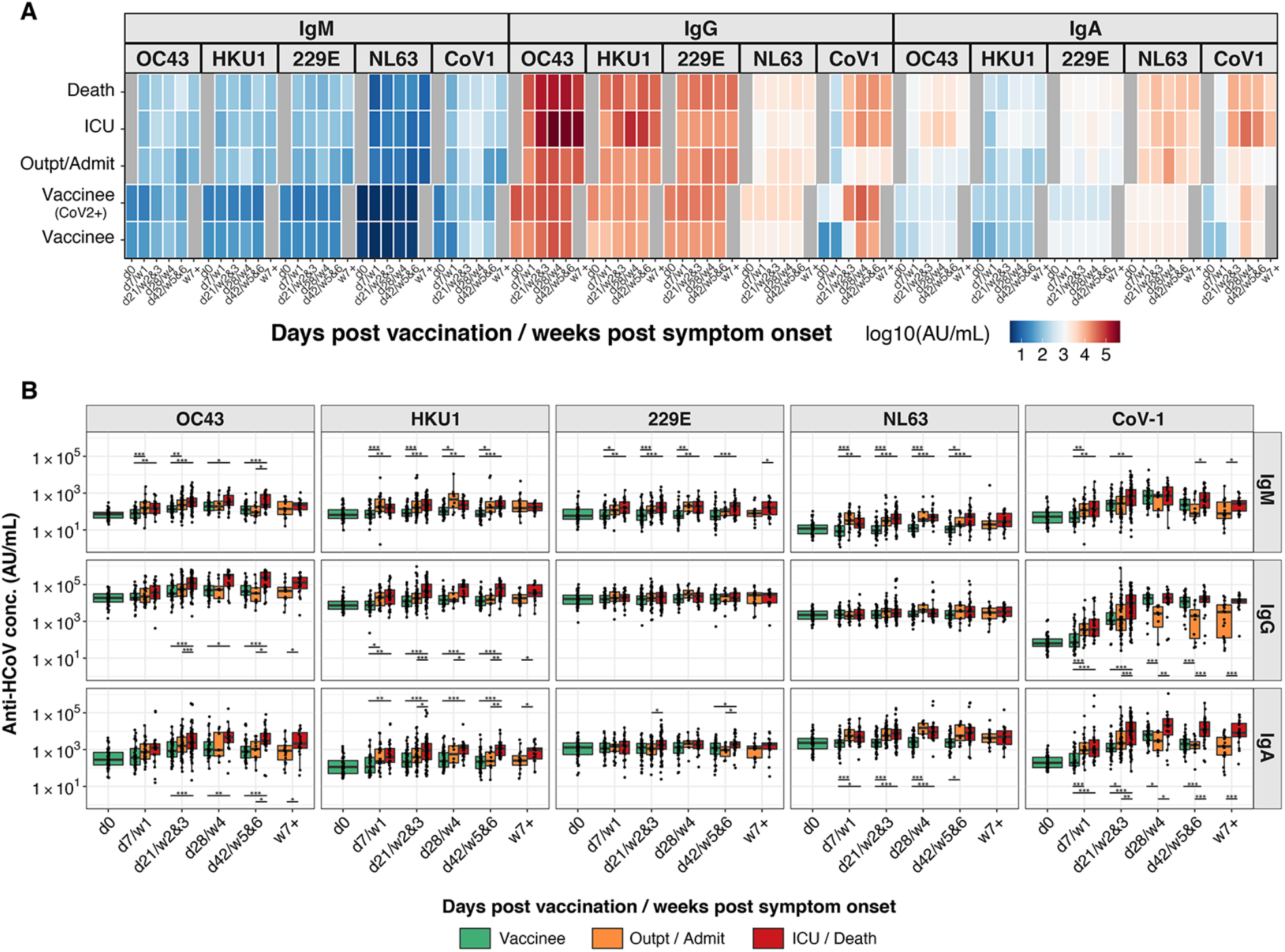
**BNT162b2 vaccination produces less broad serological responses to endemic HCoVs than SARS-CoV-2 infection.** Anti-SARS-CoV S, and anti-HCoV-OC43, -HKU1, -NL63 and −229E S IgM, IgG, and IgA antibody responses are shown for individuals who received BNT162b2 prime (day 0) and boost (day 21) vaccination doses and for COVID-19 patients. (A) Heatmap showing the development of antibody responses in longitudinal samples from vaccinees/patients collected at / during day 0, day 7 / week 1, day 21 / weeks 2&3, day 28 / week 4, day 42 / weeks 5&6, and week 7 and later after vaccination / COVID-19 symptom onset (x-axis). Individuals were classified as outpatients (Outpt) and hospital admitted patients (Admit); intensive care unit (ICU) patients, those who died from their illness (Death) and vaccinees who did (CoV-2+) or did not have a positive SARS-CoV-2 test in the past. The color scale encodes the median values of log10 MSD arbitrary unit (AU)/mL concentrations. (B) Box-whisker plots of the MSD AU/mL anti-SARS-CoV-2 Ig concentrations show the interquartile range as the box and the minimum and maximum values as the ends of the whiskers. Statistical test: pairwise Wilcoxon rank sum test with Bonferroni correction. * = P < 0.05, ** = P < 0.01, *** = P < 0.001.

### Circulating SARS-CoV-2 variants show consistent degrees of escape from polyclonal antibody responses of vaccinees and infected patients

SARS-CoV-2 variants associated with rapidly increasing case numbers have recently emerged and spread globally. Neutralizing capability of many potent anti-SARS-CoV-2 monoclonal antibodies (mAbs) against B.1.351 is reduced or abolished, and escape from recognition by vaccinee and patient plasma has been reported (Zhou et al., 2021). We analyzed and compared variant B.1.351, P.1, and B.1.1.7 S binding and S-ACE2 blocking activity of antibodies in longitudinal plasma samples from the BNT162b2 vaccinees and COVID-19 patients. Compared with Wuhan-Hu-1, antibody binding to viral variant S and RBD antigens was reduced to similar degrees in vaccinees (Figures 4A and 4B) and patients (Figure 4C), with more marked decreases for P.1 and B.1.351. Most vaccinees developed high percentages of Wuhan-Hu-1 S-ACE2 blocking activity, peaking at day 28 (7 days post-boost). A strikingly consistent hierarchy of reduction in plasma antibody binding by variant S and RBD antigens was observed among study participants, with progressively decreased binding for B.1.1.7, P.1 and B.1.351 compared to Wuhan-Hu-1 antigens. The S-ACE2 blocking antibody assay showed highly similar reductions in blocking for B.1.351 and P.1, indicating no significant difference between the effects of the K417N versus K417T RBD changes, respectively, on ACE2 blocking antibodies (Figure 4B). Two vaccinees had low RBD and S binding and S-ACE2 blocking activity for all SARS-CoV-2 variants, including Wuhan-Hu-1. Together, these data indicate that the effects of viral variants are remarkably consistent with an escape from polyclonal antibody responses elicited either by infection or BNT162b2 vaccination.

**Figure 4.**
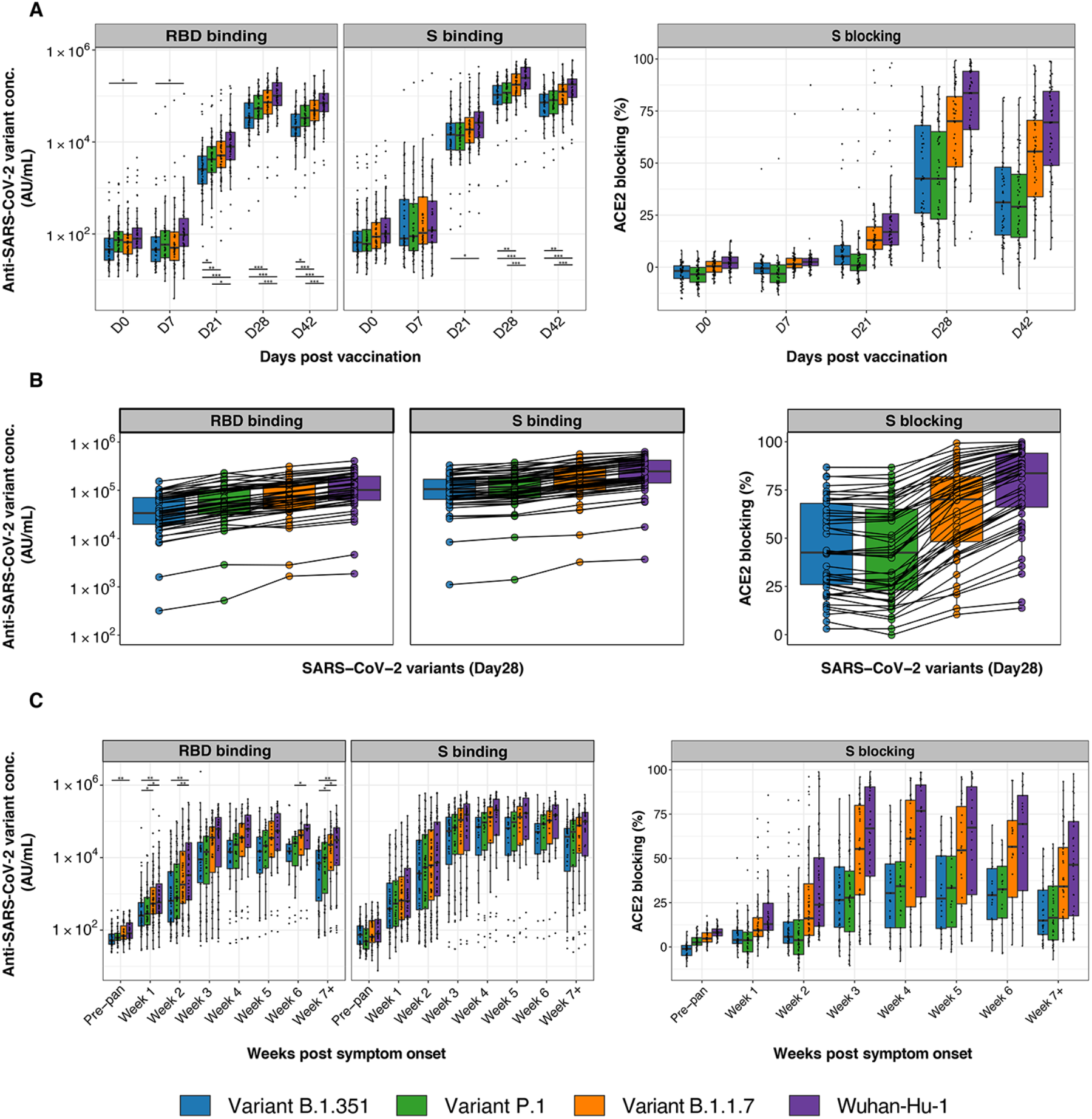
**Circulating SARS-CoV-2 variants show consistent degrees of escape from polyclonal antibody responses of vaccinees and infected patients.** Anti-SARS-CoV-2 RBD and S antibody responses are shown for Wuhan-Hu-1 and viral variants of concern (B.1.1.7, UK; P.1, Brazil; B.1.351, South Africa). Box-whisker plots of the MSD AU/mL anti-SARS-CoV-2 IgG binding concentrations and S-ACE2 blocking percentages show the interquartile range as the box and the minimum and maximum values as the ends of the whiskers. (A) Plasma samples from BNT162b2 vaccinees. (B) Comparison of antibody concentrations of BNT162b2 vaccinees on day 28 for different variants of concern. Data points for individual study participants are connected with a line. (C) Plasma samples from COVID-19 patients. Significance between groups (Wuhan-Hu-1, B.1.1.7, P.1, and B.1.351) was tested with pairwise Wilcoxon rank sum test with Bonferroni correction. * = P < 0.05, ** = P < 0.01, *** = P < 0.001.

## Discussion

One of the positive developments amid the global calamity of the SARS-CoV-2 pandemic has been the rapid design, production and deployment of remarkably effective vaccines based on lipid nanoparticle delivery of mRNA encoding the viral S antigen (Baden et al., 2021; Polack et al., 2020). Although correlates of vaccine-mediated protection are still under active study, clinical correlates and passive antibody transfer experiments in rhesus macaques support a central role for neutralizing antibodies that block the viral S protein’s interaction with the host ACE2 receptor. Such antibodies are elicited by infection as well as vaccination (Chandrashekar et al., 2020; Sahin et al., 2020; Yu et al., 2020), but the shared and divergent features of the serological responses produced in response to SARS-CoV-2 antigens in these different contexts are poorly understood.

We find that BNT162b2 vaccination produces robust IgG responses to S protein, and RBD and NTD domains that are as high as those generated in the most severely ill COVID-19 patients and follow a similar time course. Side-effects following vaccination were not associated with the magnitude of the serological response. Unlike infection, which stimulates robust but short-lived IgM and IgA responses, vaccination shows a pronounced bias for IgG production. These responses were similar across the adult age range in our study but showed slightly lower levels in individuals over 60 years of age. Candidate explanations for the relative absence of IgM and IgA responses to the vaccine are the potent effect of the lipid components of the vaccine formulation in driving early and extensive IgG class-switching, potentially as a result of the reported Th1-polarized CD4^+^ T cell responses and vigorous germinal center formation stimulated by these vaccine components (Lederer et al., 2020; Lindgren et al., 2017; Pardi et al., 2018b). Vaccinees in our study showed higher concentrations of IgG, and similar concentrations of IgA in comparison to patients with mild COVID-19 illness. In the UK-based SIREN (SARS-CoV-2 Immunity & Reinfection Evaluation) observational cohort of health care workers, estimates of SARS-CoV-2 reinfection rates compared to primary infection rates, indicated an approximately 83% reduced risk (Hall et al., 2021). The reported 95% efficacy of BNT162b2 in preventing primary infection compares favorably with this estimate and may indicate additional protection provided by the higher IgG levels produced by the vaccine.

Compared to infection, the BNT162b2 vaccine also stimulates a less broad antibody response to endemic HCoVs, despite having anti-SARS-CoV-2 and anti-SARS-CoV IgG concentrations as high as those of the most severely ill COVID-19 patients. The four endemic coronaviruses, HCoV-OC43 and HCoV-HKU1 (*Betacoronavirus*), and HCoV-NL63 and HCoV-229E (*Alphacoronavirus*) are genetically and structurally dissimilar to SARS-CoV-2, but there are regions of conservation in the S antigen (Ladner et al., 2021). Exposure to SARS-CoV-2 antigens via vaccination or infection could potentially stimulate pre-existing cross-reactive memory B cells previously generated during infections by endemic HCoVs or could generate new primary antibody responses containing cross-reactive antibodies that recognize HCoV antigens. Recent data indicate that titers of endemic HCoV-specific antibodies do not differ between SARS-CoV-2 uninfected individuals and those who become infected with SARS-CoV-2, arguing against a protective role of cross-reactive antibodies (Anderson et al., 2021). We hypothesize that differences in the inflammatory environments during SARS-CoV-2 infection compared to vaccination, and potentially the anatomical sites where the viral antigens are encountered in infection versus vaccination, may favor the more narrow SARS-CoV-2 specific antibody responses seen during BNT162b2 vaccination. Infection with SARS-CoV-2 also stimulates a broader repertoire of T cells specific for peptides from viral proteins beyond the S antigen, drawn from both memory responses to prior endemic HCoV infection and new responses to SARS-CoV-2, and therefore could provide more T cell help to a wider range of B cells in the response.

The recent emergence of SARS-CoV-2 variants with altered S protein and RBD sequences associated with immune escape has raised concern about reduced vaccine-induced immune protection. Variant B.1.1.7, first detected in September 2020 in the UK, is reported to give a 20% reduction of antibody titers in serum samples from vaccinees (Muik et al., 2021), but the ChAdOx1 vaccine based on Wuhan-Hu-1-like S antigen showed similar efficacy for earlier circulating viruses and the B.1.1.7 variant (Emary et al., 2021). Variants P.1 and B.1.351, carrying the E484K and K417 amino acid changes in RBD, further decrease recognition of antibodies stimulated by Wuhan-Hu-1-like S sequences, as exemplified by the poor efficacy of the ChAdOx1 vaccine against B.1.351 in preventing mild-to-moderate disease (Madhi et al., 2021). Here, we find that the plasmas of individuals who received prime/boost BNT162b2 vaccination, as well as COVID-19 patients, show a consistent pattern of progressively decreasing binding to the S and RBD antigens of B.1.1.7, P.1 and B.1.351, in that order. ACE2-blocking antibody activities were significantly reduced for the P.1 and B.1.351 variant in both vaccinees and COVID-19 patients collected early in the pandemic, before the spread of viral variants. These data indicate both that the proportions of polyclonal plasma antibodies targeting the epitopes affected by these viral variants are surprisingly consistent between different individuals and are comparable between infection and vaccination. The findings further suggest that susceptibility to infection by viral variants, particularly the B.1.351 and P.1 variants, is likely to be widely shared in vaccinated populations, particularly as antibody titers decrease over time. Because correlates of immunological protection are still under study, determining the extent of vulnerability to infection will require additional correlation with epidemiological surveillance for infection of vaccinated individuals over time.

Taken together, these results underscore the potent and highly targeted serological responses stimulated by the BNT162b2 mRNA vaccine, and important differences between antibody responses produced from vaccination versus infection. As investigations continue into the potential role of infection-stimulated antibodies in the lingering symptoms experienced by individuals with ‘long COVID’ syndrome, it will be important to include further evaluation of the differences in vaccination and infection serological responses. Other key questions that will require answers in the months and years ahead include the duration of effective vaccine-stimulated serological responses, and the safety and efficacy of variant-targeting vaccine boosters in previously vaccinated or infected individuals. The effectiveness of the new mRNA vaccine technologies seems likely to bring advances in the responses to other viral pathogens.

## Data Availability

All data is available in the main text or the extended materials. Code will be provided to readers upon request.

## Acknowledgments

We acknowledge Lilit Grigoryan, Yupeng Feng, and Florian Wimmers for their help in sample processing. We thank the research participants in these studies.

## Funding

This work was supported by NIH/NIAID R01AI127877 (S.D.B.), NIH/NIAID R01AI130398 (S.D.B.), NIH 1U54CA260517 (S.D.B.), NIH/NIAID HHSN272201700013C (G.B.S.), an endowment to S.D.B. from the Crown Family Foundation and an Early Postdoc.Mobility Fellowship Stipend to O.F.W. from the Swiss National Science Foundation (SNSF).

## Author Contributions

K.R., S.C.A.N., M.M.D, B.P., K.C.N., S.D.B. conceptualized and designed the study. K.R., S.C.A.N. performed the experiments. P.S.A., F.Y., R.A.H., O.F.W., A.S.L., F.G., V.M., C.L., E.H., M.S., collected and processed samples. J.L.W., J.N.W., B.A.P., G.B.S. provided reagents and samples. K.R., S.C.A.N. analyzed the data and performed statistical analyses. K.R., S.C.A.N, S.D.B. wrote the manuscript. All authors provided intellectual contributions, edited and approved the manuscript.

## Declaration of Interests

S.D.B. has consulted for Regeneron, Sanofi, and Novartis on topics unrelated to this study, and owns stock in AbCellera Biologics. K.C.N. reports grants from National Institute of Allergy and Infectious Diseases (NIAID), Food Allergy Research & Education (FARE), End Allergies Together (EAT); National Heart, Lung, and Blood Institute (NHLBI), and National Institute of Environmental Health Sciences (NIEHS). K.C.N. is Director of FARE and World Allergy Organization (WAO) Center of Excellence at Stanford; Advisor at Cour Pharmaceuticals; Cofounder of Before Brands, Alladapt, Latitude, and IgGenix; National Scientific Committee member for the Immune Tolerance Network (ITN) of NIAID; recipient of a Research Sponsorship from Nestle; Consultant and Advisory Board Member at Before Brands, Alladapt, IgGenix, NHLBI, and ProBio; and Data and Safety Monitoring Board member at NHLBI.

## Supplemental Figures

**Figure S1:**
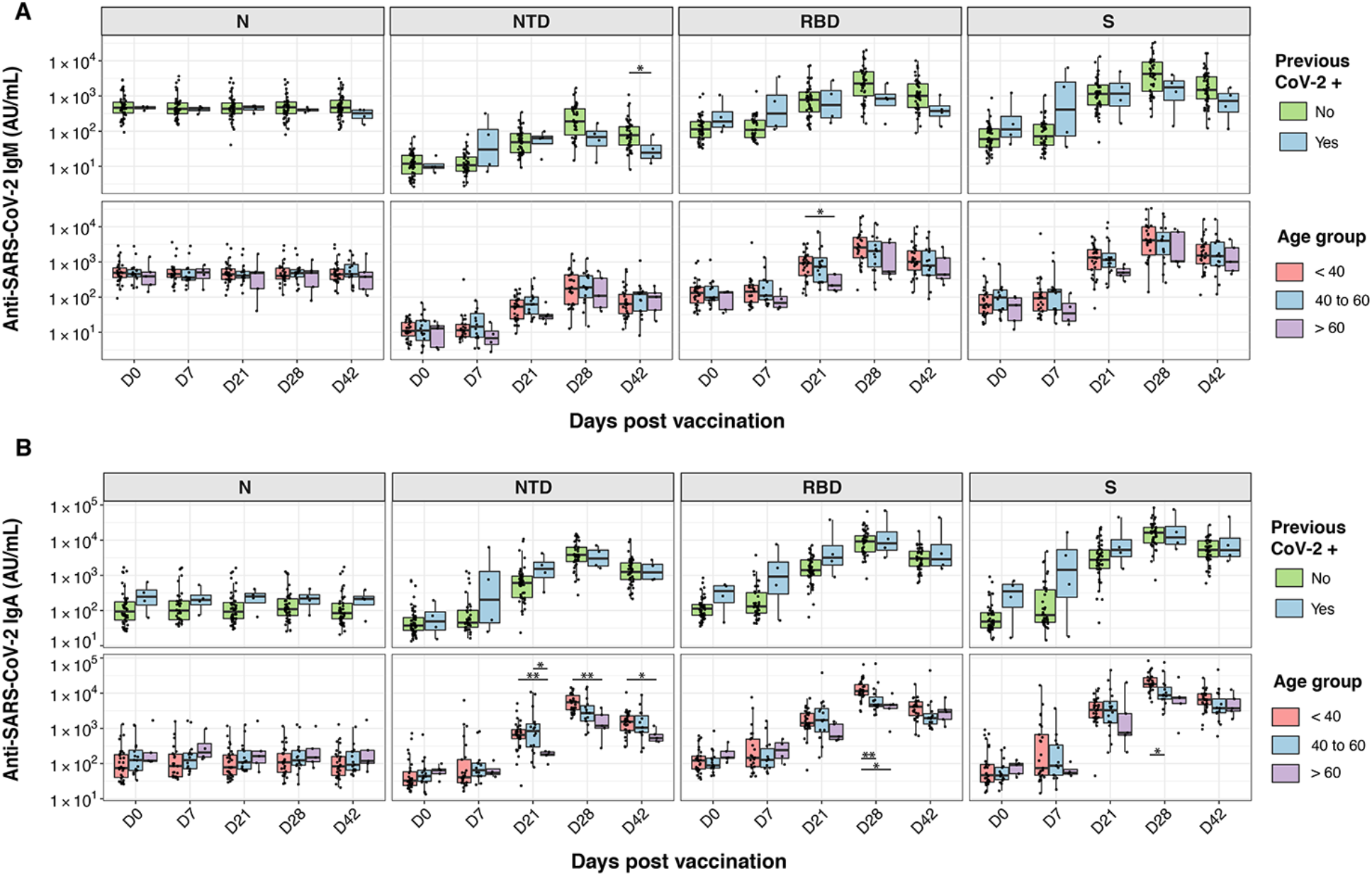
**BNT162b2 vaccination induces variable and relatively low anti-SARS-CoV-2 IgM and IgA concentrations.** Anti-SARS-CoV-2 N, NTD, RBD, and S IgM (A) and IgA (B) antibody responses are shown for 257 plasma samples from 55 individuals who received BNT162b2 prime (day 0) and boost (day 21) vaccination doses. Box-whisker plots of the MSD AU/mL anti-SARS-CoV-2 IgG concentrations show the interquartile range as the box and the minimum and maximum values as the ends of the whiskers. Statistical tests: two-sided Wilcoxon rank sum test (A and B, top panels) and pairwise Wilcoxon rank sum test with Bonferroni correction (A and B, bottom panels). * = P < 0.05, ** = P < 0.01, *** = P < 0.001.

**Figure S2:**
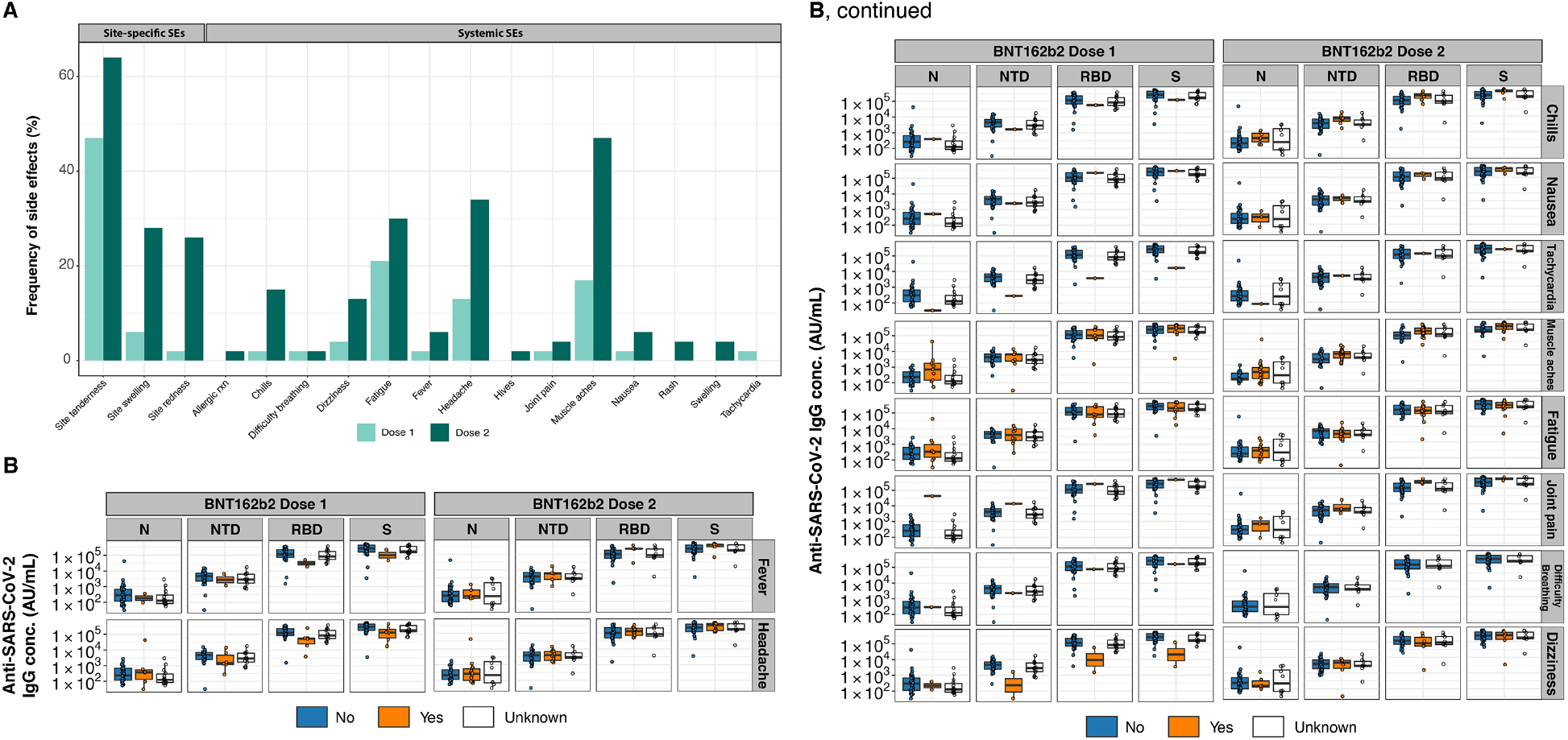
**The magnitude of antibody responses is not correlated with reported vaccine-associated side effects (SEs).** (A) Frequency of vaccine-associated side effects after the prime (light green) and boost (dark green) vaccination dose. (B) Box-whisker plots of the MSD AU/mL anti-SARS-CoV-2 IgG concentrations show the interquartile range as the box and the minimum and maximum values as the ends of the whiskers. For a given SE (horizontal panels), vaccinees were grouped according to no SE reported (“No”, colored in blue) or SE reported (“Yes”, colored in orange). Vaccinees where SEs were unknown are shown as white boxplots.

**Figure S3:**
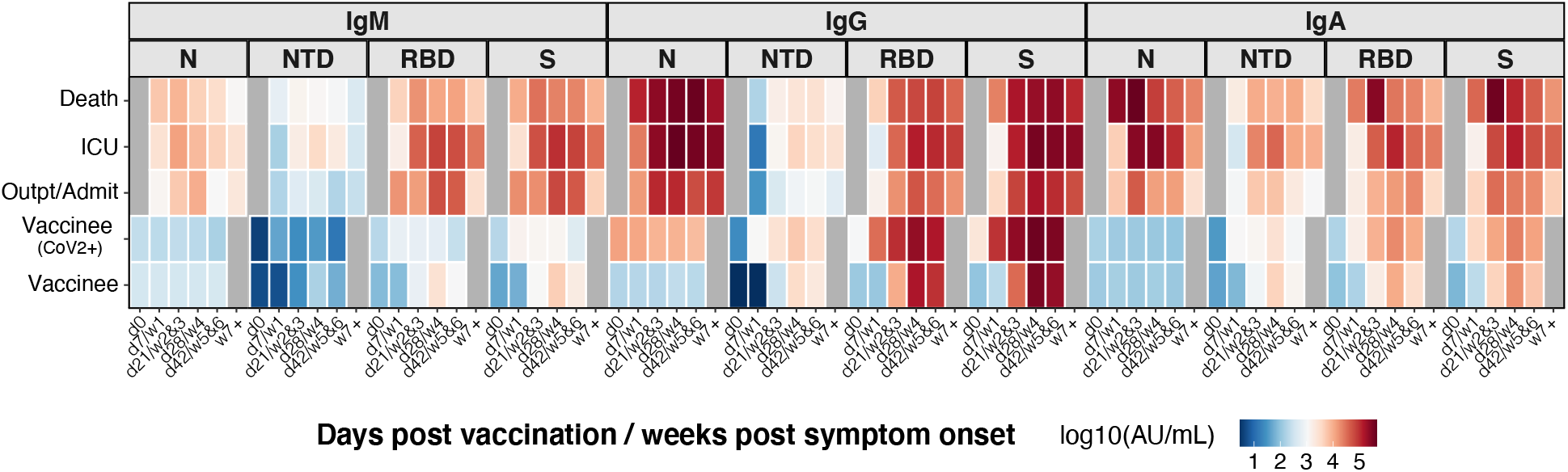
**BNT162b2 vaccination and SARS-CoV-2 infection elicit divergent Ig isotype profiles.** Anti-SARS-CoV-2 N, NTD, RBD, and S IgM, IgG, and IgA antibody responses are shown for individuals who received BNT162b2 prime (day 0) and boost (day 21) vaccination doses and for COVID-19 patients. The heatmap shows the development of antibody responses in longitudinal samples from vaccinees/patients collected at / during day 0, day 7 / week 1, day 21 / weeks 2&3, day 28 / week 4, day 42 / weeks 5&6, and week 7 and later after vaccination / COVID-19 symptom onset (x-axis). Individuals were classified as outpatients (Outpt) and hospital admitted patients (Admit); intensive care unit (ICU) patients, those who died from their illness (Death) and vaccinees who did (CoV-2+) or did not have a positive SARS-CoV-2 test in the past. **<u>Mean</u>** values (as opposed to the **<u>Median</u>** values shown in the main Figure 2) of log10 MSD arbitrary unit (AU)/mL concentrations were used to display a color code for each of the study groups.

**Figure S4:**
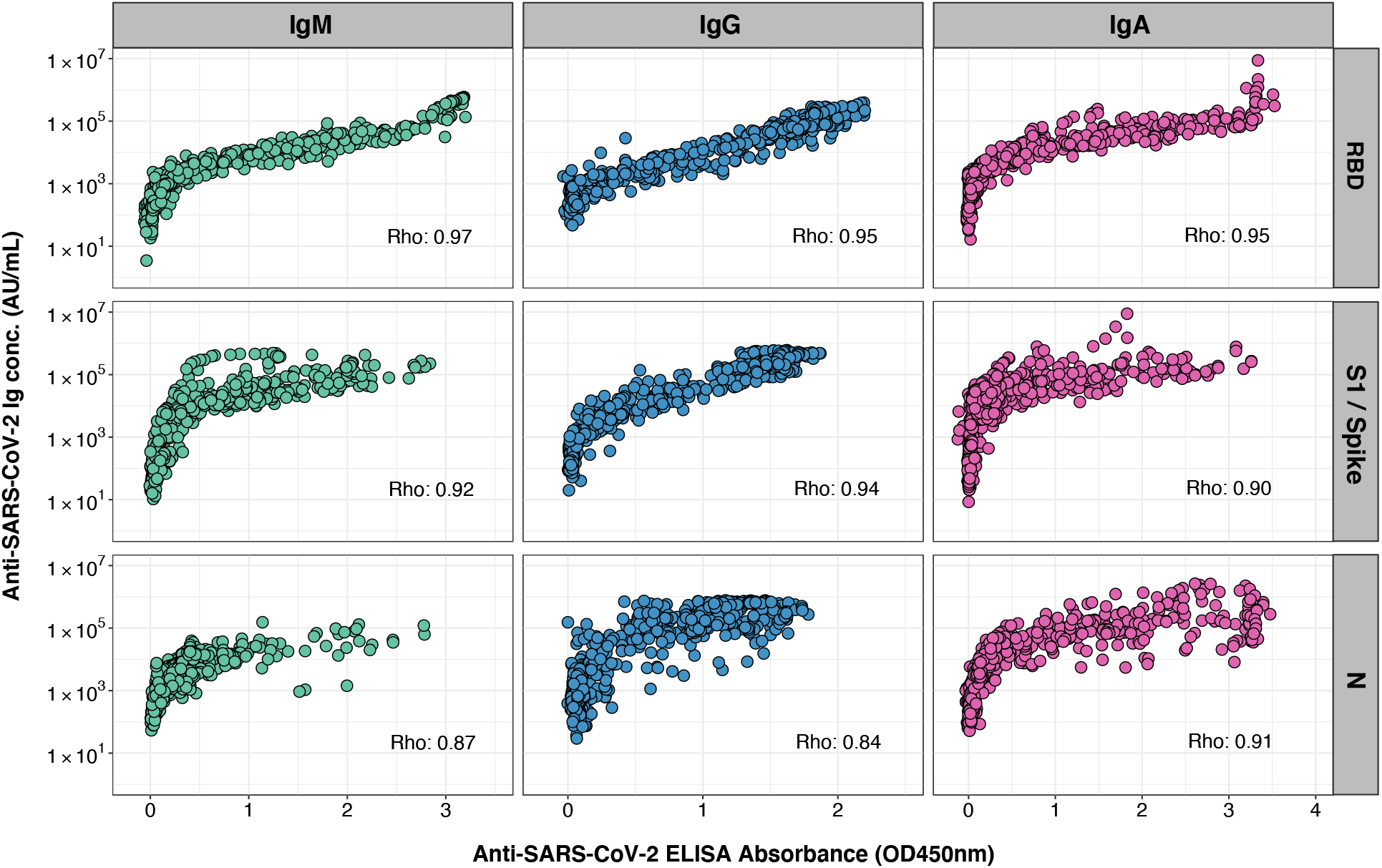
Correlation of anti-SARS-CoV-2 ELISA and ECL results. Anti-SARS-CoV-2 RBD, S1/S, and N IgM, IgG, and IgA antibody responses were measured in 530 plasma samples from 100 COVID-19 patients by ELISA and MSD ECL assays. ELISA versus MSD RBD and N assay results and ELISA S1 versus MSD S assay results were highly correlated. Spearman rank correlation (coefficient = Rho, displayed in the plot for each assay comparison) was used to assess the strength of correlation between ELISA and MSD results. Outliers for the N assays with less correlated ELISA and MSD results may be attributed to the fact that the N protein used in the ELISAs was produced in *E. coli*, whereas the MSD N protein was produced in mammalian cells, potentially causing differences in post-translational modifications and thus epitope recognition.

**Figure S5:**
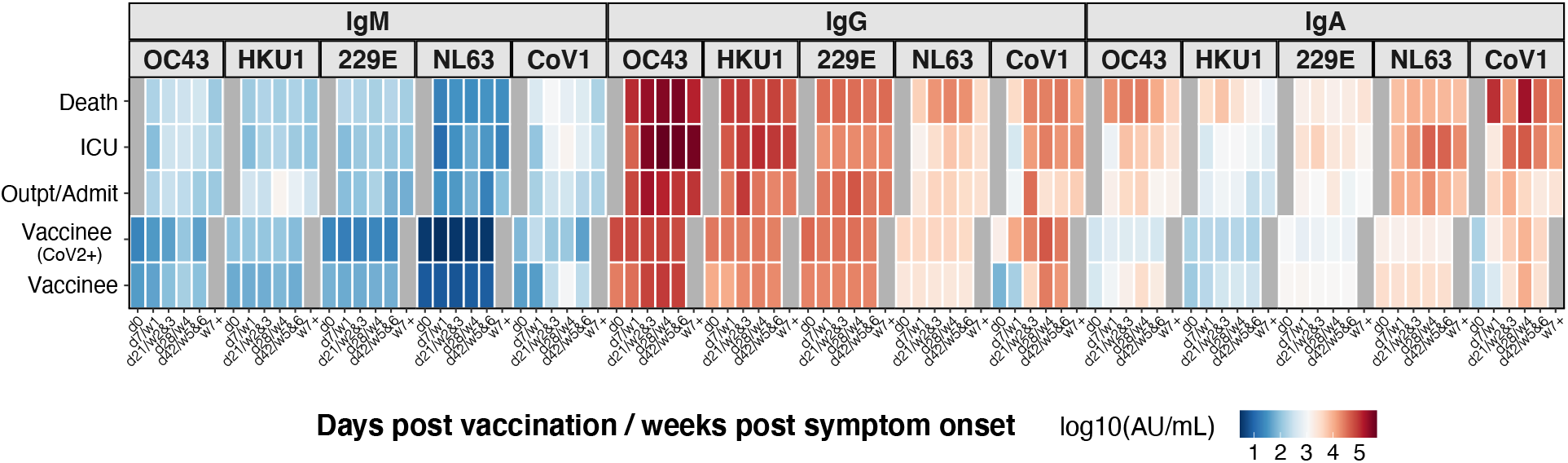
**Less broad serological responses to endemic HCoVs from BNT162b2 compared to SARS-CoV-2 infection.** Anti-SARS-CoV S, and anti-HCoV-OC43, -HKU1, -NL63 and −229E S IgM, IgG, and IgA antibody responses are shown for individuals who received BNT162b2 prime (day 0) and boost (day 21) vaccination doses and for COVID-19 patients. The heatmap shows the development of antibody responses in longitudinal samples from vaccinees/patients collected at / during day 0, day 7 / week 1, day 21 / weeks 2&3, day 28 / week 4, day 42 / weeks 5&6, and week 7 and later after vaccination / COVID-19 symptom onset (x-axis). Individuals were classified as outpatients (Outpt) and hospital admitted patients (Admit); intensive care unit (ICU) patients, those who died from their illness (Death) and vaccinees who did (CoV-2+) or did not have a positive SARS-CoV-2 test in the past. **<u>Mean</u>** values (as opposed to the **<u>Median</u>** values shown in the main Figure 3) of log10 MSD arbitrary unit (AU)/mL concentrations were used to display a color code for each of the study groups.

## STAR Methods

### RESOURCE AVAILABILITY

#### Lead Contact

Further information and requests for resources and reagents should be directed to the Lead Contact, Scott D. Boyd.

### EXPERIMENTAL MODELS AND SUBJECT DETAILS

#### Samples from BNT162b2 vaccinees

All participants in the study provided informed consent, under Stanford University Institutional Review Board approved protocol IRB-55689. To study immune responses after first and second dose vaccination with BNT162b2, we included 257 longitudinal samples from 55 vaccinees. Samples were collected on day 0 before or immediately after the first vaccination dose and individuals received their second dose on day 21. Time points in the manuscript are defined as day 7, 21, 28, and 42 and blood was drawn +/-one day from the assigned time point. Peripheral blood samples were collected in vacutainer cell preparation tubes (CPT) containing sodium citrate. After centrifugation for collection of plasma, samples were aliquoted and stored at −80°C. Demographic information for all vaccinees is provided in Table 1.

**Table 1.** Demographic characteristics of vaccine study participants.

| **Characteristics** |  | **BNT162b2 vaccinees** |
| --- | --- | --- |
|  |  | **(n = 55)** |
| **Demographic information available, n (%)** |  | 52 (95) |
| **Age, median (IQR)** |  | 36 (31.5 – 50) |
| **Sex, n (%)** | **Female** | 27 (49) |
|  | **Male** | 25 (45) |
| **Race\*, n (%)** | **Asian** | 22 (40) |
|  | **Black** | 3 (5) |
|  | **White** | 24 (44) |
| **Previous SARS-CoV-2+ test result, n (%)** |  | 4 (7) |

\* two individuals reported to be American and one individual reported to be Indian and were thus
not assigned to either of the groups.

#### Samples from COVID-19 patients

We included 530 plasma samples collected between March 2020 and August 2020 from patients who reported to Stanford Healthcare-associated clinical sites with signs and symptoms of COVID-19. SARS-CoV-2 infection was confirmed for all patients by reverse-transcription quantitative polymerase chain reaction (RT-qPCR) of nasopharyngeal swabs as described (Corman et al., 2020; Hogan et al., 2020). Data for SARS-CoV-2 serology measurements on these samples by ELISA have been reported previously (Röltgen et al., 2020). Blood samples were collected in sodium heparin-coated vacutainers. After centrifugation for collection of plasma, samples were aliquoted and stored at −80°C. The use of these samples for serology testing was approved by the Stanford University Institutional Review Board (Protocols IRB-48973 and IRB-55689).

#### Healthy human control (HHC) samples

37 plasma samples from HHCs collected before the onset of the COVID-19 pandemic were used to determine baseline antibody binding to coronavirus antigens.

### METHOD DETAILS

#### MSD ECL binding assays

Plasma samples from vaccinees and COVID-19 patients were heat-inactivated at 56°C for 30 minutes and tested with MSD ECL MULTI-SPOT 96-well plate serology panels and instrumentation according to the manufacturer’s instructions. V-PLEX Coronavirus Panel 2 kits were used to detect IgM, IgG, and IgA antibodies to SARS-CoV-2 N, S1 NTD, RBD, and S antigens and to S proteins of SARS-CoV and other HCoVs including HCoV-OC43, HCoV-HKU1, HCoV-NL63, and HCoV-229E. V-PLEX SARS-CoV-2 Panel 7 kits were used to detect IgG antibodies to different SARS-CoV-2 variant RBD and S proteins, including Wuhan-Hu-1, B.1.351, P.1, and B.1.1.7. Plasma samples were analyzed in duplicate at a 1:5’000 dilution, detected with anti-human IgM, IgG, or IgA antibodies conjugated to SULFO-TAG ECL labels and read with a MESO QuickPlex SQ 120 instrument. Each plate contained duplicates of a 7-point calibration curve with serial dilution of a reference standard, a blank well and three positive control samples. Calibration curves were used to calculate antibody unit concentrations (MSD AU/mL) by backfitting ECL signals measured for each sample to the curve.

#### MSD ECL blocking assays

Antibodies blocking the binding of SARS-CoV-2 RBD to ACE2 were detected with MSD V-PLEX SARS-CoV-2 Panel 7 (ACE2) kits according to the manufacturer’s instructions. Heat-inactivated plasma samples from vaccinees and COVID-19 patients were analyzed in duplicate at a dilution of 1:100. Samples were incubated together with human ACE2 protein conjugated with a SULFO-TAG and read with a MESO QuickPlex SQ 120 instrument. Each plate contained duplicates of a 7-point calibration curve with serial dilution of a reference standard and a blank well. Results are reported as percent inhibition calculated based on the equation ((1 – Average Sample ECL Signal / Average ECL signal of blank well) x 100).

#### ELISA testing of COVID-19 patient samples

ELISA testing of the 530 COVID-19 patient samples for the presence of antibodies to SARS-CoV-2 antigens was performed previously (Röltgen et al., 2020). To compare the performance of the MSD SARS-CoV-2 panel plates with our laboratory developed ELISAs we included results for anti-SARS-CoV-2 RBD, S1, and N IgM, IgG, and IgA ELISA testing in this study. Briefly, ELISA was performed after coating 96-well Corning Costar high binding plates (catalog no. 9018, Thermo Fisher) SARS-CoV-2 RBD, S1, or N protein in phosphate-buffered saline (PBS) at a concentration of 0.1 μg per well (0.025 μg per well for the nucleocapsid IgG assay) overnight at 4°C. On the next day, plates were blocked, and wells were then incubated with plasma samples from COVID-19 patients at a dilution of 1:100. Bound antibodies were detected with horseradish peroxidase conjugated goat anti-human IgG (γ-chain specific, catalog no. 62-8420, Thermo Fisher, 1:6,000 dilution), IgM (μ-chain specific, catalog no. A6907, Sigma, 1:6,000 dilution), or IgA (α-chain specific, catalog no. P0216, Agilent, 1:5,000 dilution). 3,3′,5,5′-Tetramethylbenzidine (TMB) substrate solution was added and the reaction was stopped after 12 min by addition of 0.16 M sulfuric acid. The optical density (OD) at 450 nanometers was measured with an EMax Plus microplate reader (Molecular Devices, San Jose, CA).

### QUANTIFICATION AND STATISTICAL ANALYSIS

Statistical tests were performed in R using base packages for statistical analysis and the ggplot2 package was used for graphics. Box-whisker plots show median (horizontal line), interquartile range (box), and 1.5 times the interquartile range (whiskers). In all analyses where statistical significance was tested, significance was defined as: ***p-value < 0.001; **p-value < 0.01; *p-value ≤ 0.05.

## Notes

### Author Declarations

All participants in the study provided informed consent, under Stanford University Institutional Review Board approved protocols IRB-55689. and IRB-48973.

